# Decreased retinal vascular complexity is an early biomarker of MI supported by a shared genetic control

**DOI:** 10.1101/2021.12.16.21267446

**Authors:** Ana Villaplana-Velasco, Justin Engelmann, Konrad Rawlik, Oriol Canela-Xandri, Claire Tochel, Frida Lona-Durazo, Muthu Rama Krishnan Mookiah, Alex Doney, Esteban J. Parra, Emanuele Trucco, Tom MacGillivray, Kristiina Rannikmae, Albert Tenesa, Erola Pairo-Castineira, Miguel O. Bernabeu

## Abstract

There is increasing evidence that the complexity of the retinal vasculature (measured as fractal dimension, D_f_) might offer earlier insights into the progression of coronary artery disease (CAD) before traditional biomarkers can be detected. This association could be partly explained by a common genetic basis; however, the genetic component of D_f_ is poorly understood. We present here a genome-wide association study (GWAS) aimed to elucidate the genetic component of D_f_ and to analyse its relationship with CAD. To this end, we obtained D_f_ from retinal fundus images and genotyping information from ∼38,000 white-British participants in the UK Biobank. We discovered 9 loci associated with D_f_, previously reported in pigmentation, retinal width and tortuosity, hypertension, and CAD studies. Significant negative genetic correlation estimates endorse the inverse relationship between D_f_ and CAD, and between D_f_ and myocardial infarction (MI), one of CAD fatal outcomes. This strong association motivated us to developing a MI predictive model combining clinical information, D_f_, a CAD polygenic risk score and using a random forest algorithm. Internal cross validation evidenced a considerable improvement in the area under the curve (AUC) of our predictive model (AUC=0.770) when comparing with an established risk model, SCORE, (AUC=0.719). Our findings shed new light on the genetic basis of D_f_, unveiling a common control with CAD, and highlights the benefits of its application in individualised MI risk prediction.

## Introduction

Coronary artery disease (CAD) remains the leading cause of death and disability worldwide^1^. Early-diagnosis and preventive therapies are essential strategies to control CAD morbidity and the mortality associated with its outcomes, such as myocardial infarction (MI). There is increasing evidence that morphological changes in the retinal vasculature, for instance in vessel width and vascular complexity, might offer insights into CAD before traditional risk factors (such as systolic blood pressure and cholesterol levels)^2,3^. Recent studies reported that a reduced degree of vascular complexity, quantified through estimates of the fractal dimension (D_f_), is found in individuals who had a higher CAD risk, independent of their age^2^. This suggests that D_f_ could be a promising non-invasive and highly accessible biomarker. Nevertheless, this finding has not translated so far to a substantial increase in prediction accuracy for major adverse cardiac events (MACE) risk, compared to models based on patient demographics and lifestyle risk factors^4,5^.

Evidence points towards coronary and retinal vessels experiencing similar pathophysiological changes at even early CAD stages^6–8^, plausibly influenced by a shared genetic basis^7,9–14^. Population-based studies have demonstrated that both tortuosity and width of arteries and veins have a moderate genetic basis^12,13^. Veluchamy *et al*. described two novel loci near the *COL4A2* and *ACTN4* genes associated with retinal tortuosity which were previously reported in genetic atrial fibrillation and CAD^11^ studies. However, the genetic component of retinal vascular complexity remains poorly understood.

We report here a genome-wide association study (GWAS) on the topic, using ∼38,000 white British participants from the UK Biobank. The aim is twofold: to comprehensively study the genetic control of D_f_ and to assess the extent of its relationship with CAD. We discovered 9 loci associated with D_f._ Two of these loci (*SLC12A9* and *RDH5* genes) were previously associated with cardiovascular risk factors and diseases^15^. Genetic correlation estimates and enrichment analysis support the shared genetic control between D_f_ and CAD, suggesting that decreasing D_f_ might be influenced by clinical CAD manifestations and, in part, by common genetic effects. Given this strong connection, we developed a model to predict incident MI cases in the UK Biobank over the 10 years succeeding the ophthalmic examination at baseline, including D_f_ and a CAD polygenic risk score (PRS_CAD_). Internal 10-fold cross validation shows a considerable performance improvement compared with the SCORE model^16^, an established CAD risk prediction score based on epidemiological variables. This enhancement can be partly explained by the additional predictive power of retinal and genetic determinants, as these respectively capture early vascular morphological abnormalities and personalised MI risk. Our findings shed new light on the genetic component of D_f_, suggesting a common genetic basis with CAD aetiology, and demonstrate its potential for individual MI risk prediction.

## Results

### Automated Quality Control and fractal dimension calculation in UK Biobank fundus images reveals interocular asymmetry in vascular complexity at an individual level

For this study, we first developed a semi-automated pipeline to segment the vasculature and select good-quality segmentations in 175,611 fundus images available in the UK Biobank (Fig. 1a) using VAMPIRE software (version 3.1, Universities of Edinburgh and Dundee)^17,18^ and a previously published fundus image classifier^19^. An image quality score (IQS) was computed as part of the classification process (see Section “Methods”). D_f_ was subsequently calculated from binary vessel maps produced automatically by VAMPIRE for ∼98,600 good quality images.

**Figure 1.**
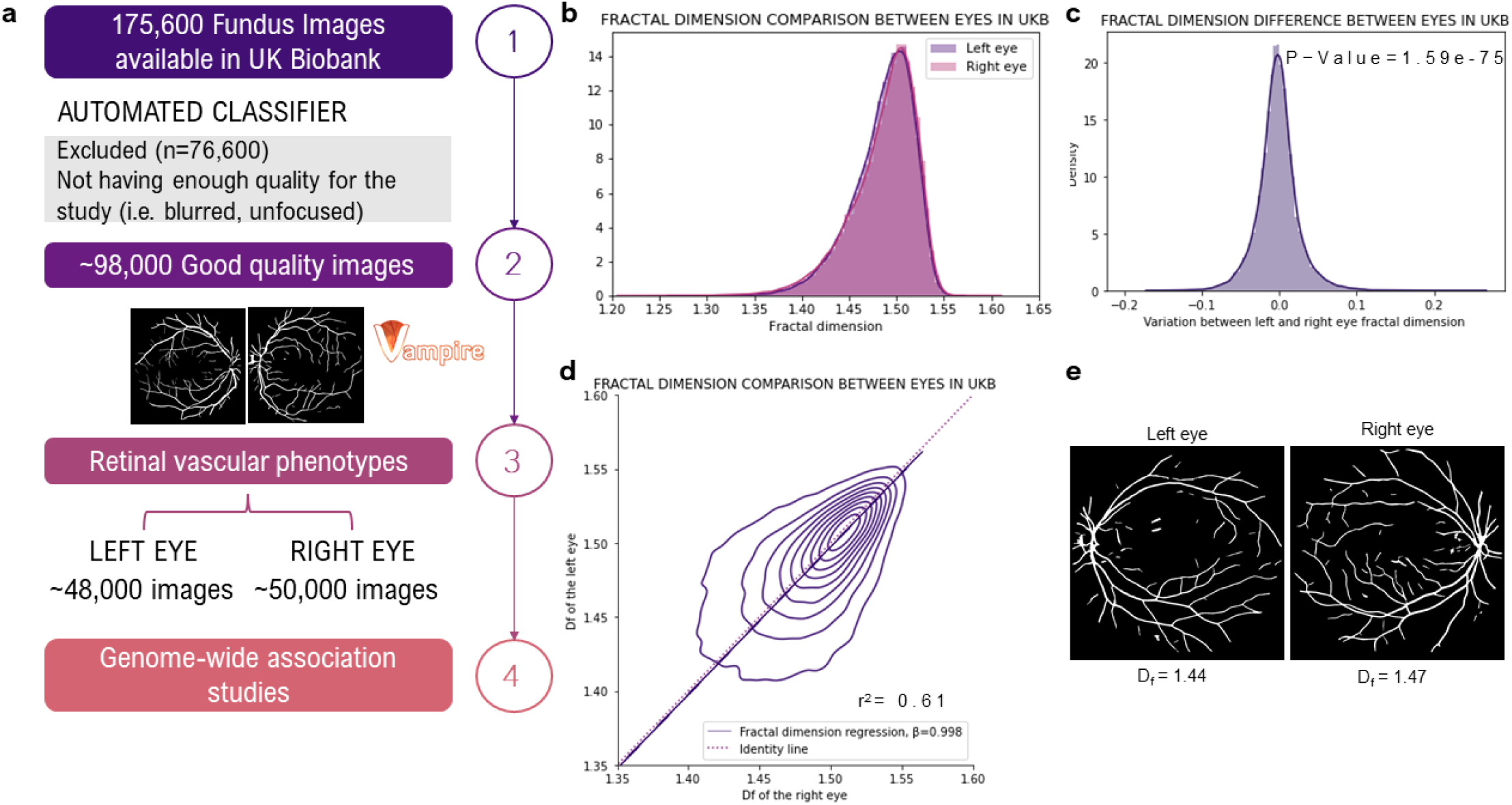
Pipeline and D_f_ characteristics. **a** Study design’s diagram describing the stepwise development of this project. **b** Left and right D_f_ histogram. **c** Individual variation distribution between left and right D_f_. **d** Overlapping left and right D_f_ histograms including the regression line. **e** Example of individual interocular asymmetry in UKBB fundus images.

We completed the, to our knowledge, largest within individual interocular D_f_ comparison (n=39,656 participants) reported so far. The population median (1.492±0.043) and D_f_ distributions appear identical between left and right eyes (Fig. 1b). However, their moderate correlation (r^2^=0.61, P-value=2·10^−16^, Fig. 1d) and the significant difference between left and right D_f_ (paired T-test P-value=1.59·10^−75^) highlight an individual interocular asymmetry (Fig. 1c), where 50% of the individuals have a right D_f_ 1 SD unit larger than their respective left D_f_. As shown in Fig. 1d, differences occur in both directions and are more pronounced when any of the D_f_ is lower than the median. To control for this individual asymmetrical effect (Fig. 1e), we performed further analysis in both eyes separately.

We next fitted univariate linear models using D_f_ as dependent variable and estimated the Pearson correlation between D_f_ and 779 UKBB binary and quantitative traits (see methods below) and IQS. Amongst these 780 variables, IQS has the strongest effect (β_right_=0.033, P-value<10^−300;^ β_left_=0.024, P-value<10^−300;^ r^2^_right_ =0.39, P-value<10^−300;^ r^2^_left_ =0.36, P-value<10^−300^). Supplementary Figure 1 illustrates this association and that a larger interocular IQS difference moderately affects D_f_ variation (β=0.014, P-value<10^− 300^). Hence, we account for IQS influence in our following analysis.

Besides IQS, 75 quantitative and 161 binary traits were significantly associated with D_f_ after Bonferroni correction^20^ (P-value<0.05/780=6.41·10^−5^). Age, sex, height, retinal disorders, smoking, hypertension, and CAD have the greatest significant effect on D_f_ in both eyes amongst all measurements (Supplementary Table 1).

### GWAS reveals nine fractal dimension loci and their association with cardiovascular risk factors

Here we present a GWAS on D_f_. This was completed with 38,811 and 38,017 unrelated white-British UK Biobank participants that had a right and left D_f_ measure, respectively. After QC (see Section “Methods”), there were 9,275,849 imputed SNPs with HWE>10^−6^, MAF>5·10^−3^, a call rate>0.9, and an imputation score>0.9. The GWAS model included hair and skin colour to control for spurious associations given the influence of eye and skin colour on fundus colour^21,22^. Hair colour replaced eye colour because the latter is not recorded during UKBB assessments. In addition, we completed a supplementary GWAS including an eye colour PRS based on the study by Lona-Durazo *et al*.^23^, which indicated no eye colour effect in our GWAS results (see Section “Methods”). The quantile-quantile plot of both GWASs indicated an adequate control of the genomic inflation in our analysis (λ_GC =_ 1.065 and λ_GC =_ 1.067 in the right and left eye, respectively, see Supplementary Fig. 2). Fig. 2c illustrates the SNPs effects comparison between eyes GWAS studies, highlighting analogous results.

**Figure 2.**
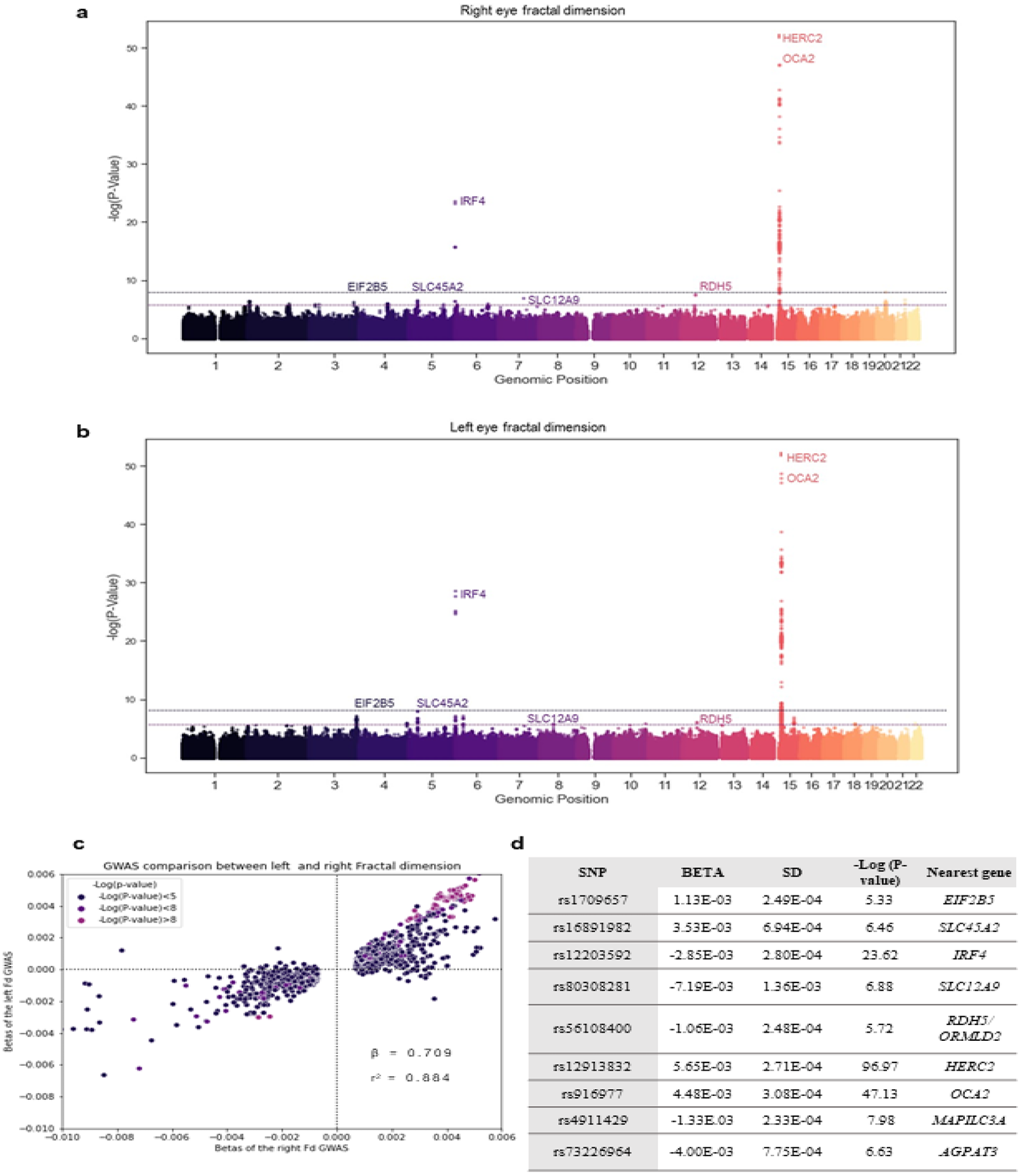
GWAS of both eyes’ D_f_. Manhattan plot of **a** right and **b** left D_f_. Points are truncated at –log10(P)=50 for clarity. **c** Comparison of the genetic variant effects between left and right D_f_ results. Colour depth indicates the significance of each variant (navy, violet, and purple for non-significant, close to genome-wide significance and significant, respectively). Genetic variants included are truncated at a minimum –log10(p) = 3 for clarity. **d** Summary statistics of D_f_ -associated SNPs and its nearest located gene.

The strongest associations are located around *OCA2* (rs916977, P-value=7.41·10^−48^) and *HERC2* (rs12913832, P-value=2.16·10^−96^) genes in chromosome 15 (Fig. 2a, Fig. 2b and Supplementary Table 2). We observed another significant association near the *IRF4* gene (rs12203592, P-value=6.59·10^−24^). Phenome-wide association studies (PheWAS), using GeneATLAS^24^ and GWASCatalog^25^, have commonly reported these SNPs in skin, hair, and eye colour analyses. Recent ocular studies demonstrated their implication in lens disorders, cataract, glaucoma, visual acuity, and retinal venular and arteriolar width and tortuosity. Additionally, Zekavat et al.^26^ recently published a paper completing a D_f_ GWAS with UKBB which reported similar results to our GWAS. However, we couldn’t make a complete comparison between studies as they didn’t make the summary statistics available yet. The comparison between reported variants of ^26^ is in Supplementary Table 3.

In addition to these 3 genetic variants, we found 6 SNPs with a P-value<10^−05^ which did not reach genomic-wide significance (Fig. 2d). The majority of these SNPs are located around genes previously reported in pigmentation analyses: *SLC45A2, SLC12A9, RDH5/ORMDL2* and *MAPILC3A*, whereas those near *EIF2B5* and *AGPAT3* genes were described in blood content and inflammation GWASs. Those SNPs located close to the *SLC12A9, RDH5/ORMLD2* and *AGPAT3* genes have also a strong effect in multiple ocular traits and diseases (such as macular thickness and retinal detachment), hypertension, and arterial disorders.

The SNP heritability (h^2^_SNP_) of the left and right D_f_ estimate are respectively 0.09 (0.015) and 0.10 (0.014). These h^2^_SNP_ magnitude is in line with previous results from retinal vascular tortuosity ^11,14^, retinal width^12^, and the recently published D_f_ ^26^ GWAS.

We completed additional D_f_ GWAS using independent UKBB participants with white European (n_left_=4340 and n_right_=4288), Asian (n_left_=562 and n_right_=568), and African (n_left_=498 and n_right_=509) ancestry. Only the GWAS including white European ancestry replicated the effect of the strongest associations (P-value<0.05/11=0.0045), which can be explained by the considerably larger number of participants in this analysis when compared with Asian and African ancestries. Little heterogeneity and forest plots of D_f_ loci indicate that multiple significant genetic variants (rs16891982, rs12203592, rs12913832 and rs31381412) have similar effect across Asian, African, white-European, and white-British ancestries (Supplementary Fig 3).

### Genetic correlation estimates and functional analysis indicate shared genetic control between fractal dimension and coronary artery disease

To assess the link between D_f_ and CAD risk factors and outcomes, we calculated their genome-wide genetic correlation using LD score regression (LDSC)^27^. Genetic correlation estimates (r^2^_g_) suggest a negative correlation between D_f_ and hypertension (r^2^_g_ =-0.30, P-value=4.52·10^−06^), acute MI (r^2^_g_ =-0.16, P-value=0.03), and CAD (r^2^_g_ =-0.18, P-value=0.025) (Table 2). Atherosclerosis and D_f_ also have a negative r^2^_g_, but significance is above the commonly accepted significance threshold (P-value<0.05). All these estimates agree in direction with phenotypic correlations (see Supplementary Table 1) and published studies, which reported that retinal D_f_ decreases as people develop these conditions^2,3,6,28^. Therefore, our results suggest that that these correlations of phenotypes are partly explained by its shared genetic basis.

**Table 2.** Genetic correlation estimates and significance (P-value) between D_f_ and associated cardiovascular events.

| Correlated trait | LEFT EYE |  |  | RIGHT EYE |  |  |
| --- | --- | --- | --- | --- | --- | --- |
| | $r_g^2$ | SE | P value | $r_g^2$ | SE | P value |
| Hypertension | -0.2229 | 0.0534 | 3.0261e-05 | -0.3020 | 0.0659 | 4.52e-06 |
| Acute myocardial infarction | -0.1717 | 0.0809 | 0.0801 | -0.1585 | 0.1075 | 0.0308 |
| Self reported acute myocardial infarction | -0.2071 | 0.0754 | 0.006 | -0.2663 | 0.0982 | 0.0067 |
| Coronary artery disease | -0.2214 | 0.0591 | 1.7854e-04 | -0.1776 | 0.0795 | 0.025 |
| Atherosclerosis | -0.3585 | 0.1819 | 0.084 | -0.2668 | 0.2100 | 0.051 |
| Right Fractal dimension | 0.9468 | 0.0962 | 1.75E-26 | - |  | - |

Moreover, we estimated the r^2^_g_ between pigmentation traits and D_f_ to examine the similarities among their genetic basis (Supplementary Fig 4). Although the estimates are non-significant (r^2^_g_ =-0.0751, P-value=0.64), genomic regions around common genetic variants in these traits might have a significant value. We did not further pursue this hypothesis experimentally.

We investigated possible causal relationships between CAD, hypertension, MI and D_f_. We found evidence of horizontal pleiotropy on the loci of interest (P=0.0056), suggesting that the link between D_f_ and such cardiovascular events may not be causal (Supplementary table 4).

FUMA functional analysis showed that D_f_ loci have a significant enrichment in melanin synthesis and pigmentation diseases as well as inflammation pathways. In addition, plasma activator-inhibitor levels type I, age-related macular degeneration, heart rate response and red blood cell count share significant overlapping gene sets with D_f_ (Supplementary Fig. 5).

### Fractal dimension improves prediction of incident myocardial infarction in UK Biobank cases

Given our findings, we hypothesized that D_f_ and PRS_CAD_ can provide additional information for MI risk estimation at an individual patient level. We thus developed a model to predict incident cases of MI over the 10 years succeeding the ophthalmic examination at baseline (Fig. 3a). Briefly, the model includes PRS_CAD_ derived from meta-analysis completed by the CARDIoGRAMplusC4D Consortium^15^, clinical variables from an established CAD risk assessment strategy named SCORE^16^ (age, sex, SBP and BMI), and the D_f_ of both eyes.

**Figure 3.**
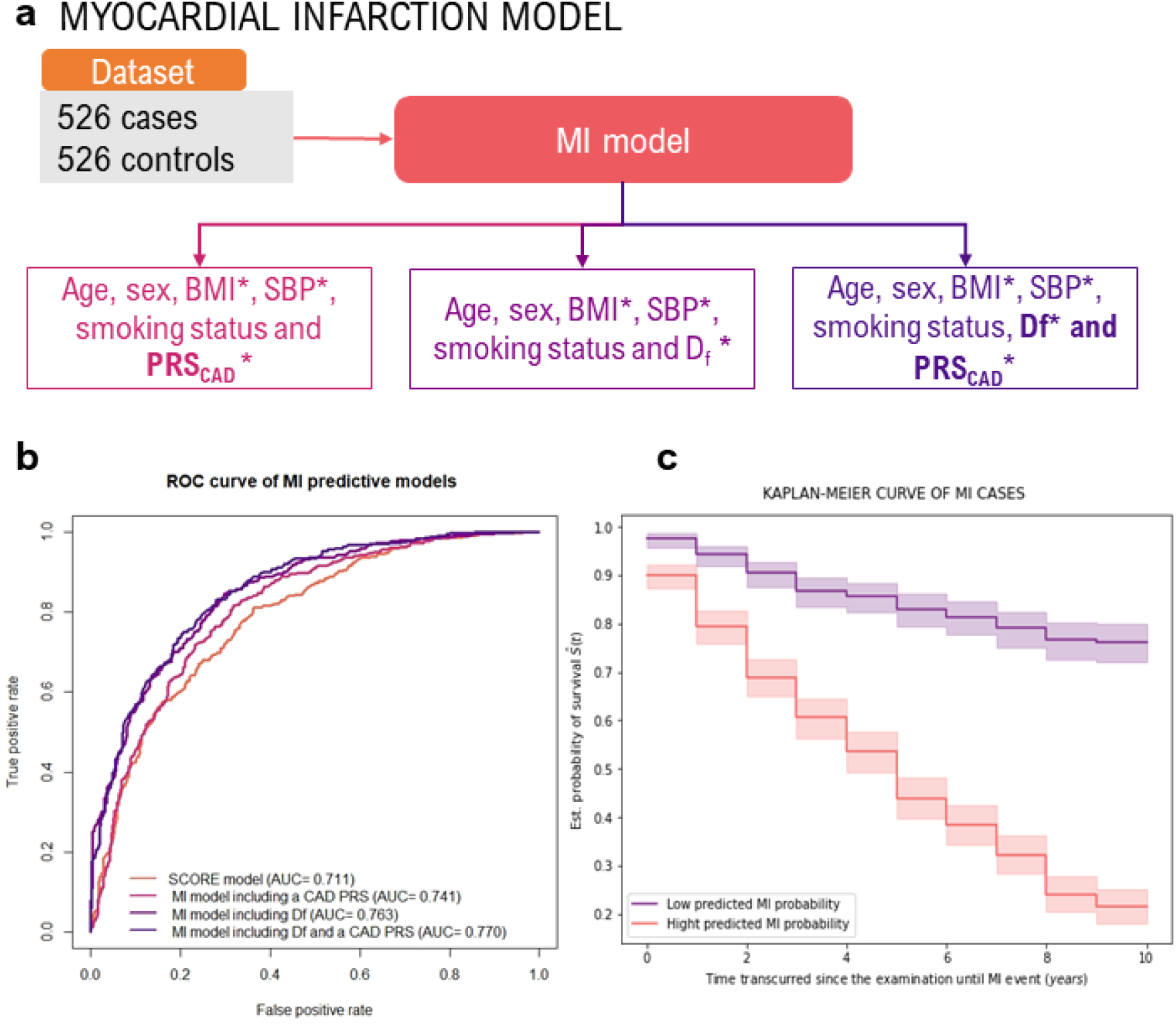
Development and performance of MI predictive models. **a** Diagram illustrating the development of our MI model. **b** ROC curve of MI predictive models. **c** Kaplan-Meier curve of incident MI cases separated by predicted MI probability.* D_f_: fractal dimension; PRS_CAD_: CAD polygenic risk score; BMI: Body-mass index; SBP: Systolic blood pressure.

The MI model was trained with the 526 individuals who experienced an MI event after their UKBB ophthalmic examination. We created an age-matched control group with an equal number of individuals and no underlying MI and CAD (Supplementary Table 5). The mean age and its SD in case and control group is respectively 57.31±6.47 and 54.21±7.84 years. We chose the random forest classifier (RFC) as this method allows one to model non-linear associations with the outcome and interactions between the predictor variables, which might boost the prediction while being interpretable^29,30^. We also considered model versions excluding either PRS_CAD_ or D_f_. As a baseline for comparison, we retrained the original SCORE model^16^ on the same dataset. Internal 10-fold cross validation (FCV) indicates that our models dominate the ROC curve of the SCORE model, achieving a greater precision, recall, and AUC (Fig 3b and Table 3). Amongst our considered models, only the one including PRS_CAD_ (AUC=0.741±0.001) yielded an AUC significantly different from the model introducing D_f_ (AUC=0.763±0.001), and the one combining both D_f_ and PRS_CAD_ (AUC=0.770±0.001) (Table 3 and Supplementary Table 6).

**Table 3.** Internal 10-fold cross validation of MI models evaluated with precision, recall and AUC.* AUC estimates significantly different (Wilcoxson signed-rank test P-value<0.005) from the ones obtained with the SCORE model. The obtained Wilcoxon signed-rank P-value for each model comparison is included in Supplementary Table 6.

| Model | MI MODEL |  |  |
| --- | --- | --- | --- |
| | Precision ± SE | Recall ± SE | AUC ± SE |
| SCORE model<sup>16</sup> | 0.716 ± 0.0025 | 0.725 ± 0.0038 | 0.719 ± 0.0021 |
| Random Forest including $D_f$ | **0.756 ± 0.0012** | **0.778 ± 0.0019** | **0.763 ± 0.0011\*** |
| Random Forest including PRS<sub>CAD</sub> | **0.735 ± 0.001** | **0.756 ± 0.0021** | **0.741 ± 0.0012\*** |
| Random Forest including $D_f$ + PRS<sub>CAD</sub> | **0.763 ± 0.0016** | **0.788 ± 0.0014** | **0.770 ± 0.0013\*** |

Next, we investigated potential survival rate differences between low and high MI risk groups. These groups were defined by the predictions obtained with our top-performing MI model and by subsequently separating these with a probability threshold of 0.5 (high MI-risk>0.5 and low MI-risk=<0.5). The Kaplan-Meier curve (Fig. 3c) illustrates a significant divergence between these groups (Log-rank test P=3.52·10^−30^), which can be explained by the pronounced decrease in survival during the first 4 years in the high MI risk group.

Finally, we performed an ablation study to understand the origin of the performance improvement in our new model. Briefly, we evaluated the performance of all possible variations between SCORE and the top-performing model (see Section “Methods”). This assessment revealed three key contributors to the reported improvement: the use of quantitative variables, the introduction of PRS_CAD_ and D_f_, and the use of a random forest classifier. An extended discussion can be found in the Supplementary Material and Supplementary Table 7. The added predictive value of D_f_ is supported by the RFC development analysis which reveals that age, BMI and D_f_ are the most important features in its architecture (Supplementary Fig.6). PRS_CAD_ is also determinant to the model’s development as its RFC importance is equivalent to SBP and smoking taken together, which in line with recently published results^8^. Additional assessments suggest that the replacement of D_f_ measures with D_f_ adjusted by IQS or just the introduction of one-eye D_f_ measurements in our MI model yield a performance comparable with the aforementioned ones (Supplementary Table 8).

## Discussion

This work provides a comprehensive examination of the genetic basis of D_f_ and unveiled a shared genetic basis with cardiovascular risk factors and CAD outcomes. Given the strong D_f_ and MI connection, we presented a predictive model for MI based on a random forest algorithm which includes D_f_ and a CAD polygenic risk score. This novel model improves MI individual risk prediction compared to state-of-the-art approaches, demonstrating the additional predictive power of these complementary traits to early identify high risk groups.

We identified an individual interocular D_f_ asymmetry in UKBB that led us to perform most of the analyses in both eyes separately. This finding is in line with published studies which reported lateral asymmetry in D_f_, tortuosity, and retinal width^31^. We observed that this asymmetry is more pronounced when one of the two eyes has a D_f_ below the population median. Interestingly, the regression coefficients and the Pearson’s correlation estimates between D_f_ and UKBB traits, and the genetic findings are equivalent in both eyes independently, suggesting that the asymmetrical effect has a negligible influence at a population level. A quantitative assessment of the asymmetry of retinal vascular measurements between eyes seems crucial for studies on retinal vascular biomarkers, often conducted on a single eye, and require further work.

We found that age, sex, smoking, and developing ocular and cardiovascular diseases have a significant effect on D_f_, agreeing with studies reporting that D_f_ decreases with age or by developing these conditions^2,6,9,13^. Interestingly, IQS has the strongest effect on this trait. To overcome quality imaging differences, various studies insist on the importance of assessing quantitatively image quality, especially in large cohorts analysed automatically^32^. In our case, IQS is computed from the binary vessel map and encapsulates the vessels segmentation’s sharpness and connectivity, which are key features frequently used ^32,33^ to compute vascular branching complexity.

We discovered 9 loci associated with D_f_ with similar effect across European, Asian, and African UKBB participants. Most of these genetic variants are relevant to multiple traits and diseases; for instance, the one located near *HERC2* has been previously associated with hair^34^, skin^35^, and eye colour^36^; but recent studies also suggest a strong effect in AMD^37^, glaucoma^38^, intraocular pressure^39^, visual acuity^40^, retinal arterial width^41^ and arterial and venular retinal tortuosity^11,14^. Another interesting associated SNP is the one near the *SLC12A9* gene as it has been reported in pigmentation^34–36^, mean arterial pressure^42^, and resting heart rate^43^ GWAS. We found a significant negative r^2^_g_ estimates between D_f_ and hypertension, CAD, and MI. The direction of these estimates agrees with their phenotypical correlations and published papers^6^, suggesting a partial explanation by their common genetic basis. This finding agrees with three aforementioned studies^11,14,41^ which identified novel retinal width and tortuosity loci associated with CAD but did not estimate a genetic correlation between retinal phenotypes and CAD. In summary, our analysis characterises the genetic architecture of D_f_ and highlights a common genetic basis with cardiovascular diseases, which partly explains the inverse association between D_f_ and coronary vessels dysfunction. Future analyses are needed to characterise the genetic expression and regulation of retinal tissue to better understand this shared genetic basis with MI and CAD.

The potential of the retinal vasculature for stratifying risk of Major Adverse Cardiac Events (MACE) has already been assessed in diabetic^5^ and non-diabetic^4,44^ individuals. Several predictive models have included retinal traits, either in a semantic^5^ or a non-semantic construction^4^, but reported very modest improvements in terms of AUC compared to the established risk estimation strategies based on epidemiological variables (e.g., 0.73 vs 0.72 in ^4^). This discrepancy with our results might be attributed to the different clinical definition of MACE, comprising normally a heterogeneous group of cardiovascular events where some of which might be not well captured in secondary care data. This situation reduces the model’s statistical power as there might be an overlap in case-controls groups. In the case of diabetic population studies, both cases and controls also have comorbidities directly affecting the architecture of the retinal vasculature that might reduce predictive power for MACE risk. In this work, we focused on MI events and considered available ICD10 guidelines and UKBB validation reports of MI data to characterize cases, achieving the maximum possible statistical power.

Recent papers have addressed the additional predictive value of a CAD PRS in MACE and CAD risk stratification^5,8,45–47^. These approaches, although mainly developed in European populations, achieve a better identification of high-risk MI individuals than those strategies based only on epidemiological variables^8,45–47^. Given this promising finding and the observed shared genetic basis between D_f_ and MI, we examined the effect of both retinal and genetic determinants on MI event risk stratification. We found that adequate clinical phenotyping is key to our models’ performance, but, as shown by our ablation study, the choice of the random forest algorithm, the use of continuous variables and the introduction of D_f_ and PRS_CAD_ in the model all independently improve traditional individual MI risk predictions. Additionally, the model including these three modifications achieves the greatest performance. We therefore speculate that D_f_ provides an early indication of coronary abnormalities not captured in clinical variables and that PRS accounts for the individual protective/risk-conferring effect on the genetic architecture of the disease. Hence, the proposed model has potential, as illustrated in the Kaplan-Meier analysis, to stratify patients by MI risk. This could allow for targeted preventive efforts, like the administration of cholesterol-lowering treatments.

Our work has multiple limitations. Firstly, there are only 526 MI cases with a good-quality fundus image taken in UKBB. Higher numbers of such participants would allow us to train and evaluate our models more robustly. Secondly, PRS_CAD_ is obtained from a meta-analysis of primarily European ancestry. This restricts the application of the predictive strategy to European samples. It is of utmost importance to complete GWAS in non-European populations to provide input for PRS estimations and to be included in its applications. Thirdly, the stability of numerical estimates of the fractal dimension is the object of a continuing debate in the retinal image analysis community^48–50^. Finally, there is little information about the genetic expression profiles and the regulation mechanisms of retinal and ocular tissues in public databases. This might be influenced by the minority of studies across these tissues and the complicated protocols to extract and characterise them.

In conclusion, our study contributes to a growing body of evidence showing associations between abnormal morphologic characteristics in coronary vessels and retinal vascular remodelling. In particular, we characterised retinal fractal dimension at a population level and uncovered the association of 9 novel loci and a shared genetic basis with CAD. Remarkably, our MI model improved the stratification of the high-risk population. This is of great interest as it discloses a promising holistic strategy that can prevent MI incidence and triage those with an elevated MI hazard. This study ultimately sheds new light on the value of easily accessible vascular imaging phenotypes and their promising application in personalised medicine.

## Materials and methods

### UK Biobank

UK Biobank (https://www.ukbiobank.ac.uk/) is a large multi-site cohort study that consist of 502,655 individuals aged between 40 and 69 years at baseline, recruited from 22 centres across the UK during 2006-2010. The study was approved by the National Research Ethics Committee, reference 11/NW/0382, and informed consent was obtained from all participants as part of the recruitment and assessment process. From these, a baseline questionnaire, physical measurements, and biological samples were undertaken for each participant. Ophthalmic examination was not included in the original baseline assessment and was introduced as an enhancement in 6 UKBB centres across the UK. This examination consisted of capturing paired retinal fundus with a 45º primary field of view and optical coherence tomography images obtained with Topcon 3D OCT-1000 MKII (Topcon Corporation). This project was completed using fundus images collected in the first and the repeated ophthalmic examination which took place in 2012 and 2013. It includes 175,709 fundus images (87,552 left and 88,157 from the right) from 67,725 participants.

### Image classification

Image quality was not reported in the cohort, and was found wanting for the purpose of automatic analysis in the first study of this kind^51^. A previous study defined an automated classifier for this dataset using three imaging features following vessels segmentation: white pixel ratio (WPR), largest connected component ratio (LCCR) and number of connected components (NCC) on a support vector machine (SVM) classifier^19^. We reproduced this classifier using a data subset of 448 random fundus images and VAMPIRE 3.1 software running in MATLAB 2018a^17,18^. The software performs automatic detection of the retinal vasculature, creating a binary vessel map for each image. A.V.V. manually classified the quality of these images based on the connectivity and the sharpness of the binary vessel map, and the lack of imaging artefacts. Manual classification was repeated 2 times using the same random subset of 100 images and the intra-classifier agreement coefficient was 0.897. This dataset was subsequently split in a training (n=278) and validation (n=170) sets. Both data subsets included an even number of manually classified good and bad quality images. We obtained a precision of 0.95, and a recall of 0.87, agreeing with the original study.

The classifier found 98,603 images with good quality from a total of 175,709 fundus images, of which 49,903 were from the right eye and 48,700 from the left eye. These images derived from ∼45,000 participants with different ancestries and included individuals with both or one eye examined at least one time. In the case of those participants that had two good quality images from one eye, following analyses are completed using the images obtained at the first examination.

Beside classification, the classifier returns an imaging quality score (IQS) based on the distance of an image from the classification boundary computed at the training phase of the SVM. We retrieved IQS using the score parameter in the prediction function running in MATLAB 2018a. We thus quantify individually the reliability of each image being classified as bad and good image.

### Calculating fractal dimension

Retinal fractal dimension, D_f_, was computed from the binarized good-quality images using VAMPIRE software based on the multifractal analysis method^52^. This process was parallelised using 12 cores and 10GB per core.

### Statistical analysis

To compare left and right D_f_ values we used participants who had both eyes scanned at the same UKBB examination and whose images were classified as good quality. 39,659 participants met these criteria. Both D_f_ distributions were compared using a paired T-Test and by estimating the Pearson correlation with the SciPy package in python 3 .We also fitted a linear regression using respectively left and right D_f_ as dependent and independent variables.

We estimated the Pearson correlation and the effect of 779 UKBB traits on D_f_ by fitting univariate linear regressions with each variable and using D_f_ as dependent variable. This included 121 quantitative variables (such as age, height, and BMI) and 658 binary variables (such as sex, diagnosed with myopia, and diagnosed with hypertension) which were extracted as reported elsewhere in ^24^. The effect of IQS was also analysed following this approach. In addition, we evaluated the IQS difference effect on D_f_ variability by fitting univariate linear regression using participants who had a good-quality image of both eyes scanned at the same UKBB examination. These analyses was completed using SciPy in python 3. Allied graphs were created using matplotib and seaborn graphical packages in python 3.

### Genome-wide association studies

We included 38,811 and 38,017 individuals in the right and left GWAS, respectively, with a self-reported and genotyped confirmed unrelated white-British ancestry^53^. Variants included were autosomal SNPs present in the genotyping arrays employed by UKBB and from the UKBB imputation panel with HWE>10^−6^, MAF>5·10^−3^, call rate>0.9 in unrelated white British individuals and imputation score>0.9 in the latest ones. The number of total SNPs analysed after quality control was 9,275,849.

Following genotype-level QC, a linear regression model was used to analyse the association of each SNP genotype with D_f_ using PLINK v2.0. We assumed an additive genetic model, adjusting for age at examination, sex, IQS, assessment centre, the first 10 genomic principal components and genotyping batch. Additionally, we included hair and skin colour as covariates to control for the influence of skin and eye colour on the fundus image colour, which can affect image segmentation and D_f_ calculation. Hair colour replaced eye colour as the latter was not recorded during UKBB assessments, and it has a similar genetic control to eye pigmentation. Besides, we performed an additional GWAS including a polygenic risk score (PRS) for eye colour to assess its influence on our GWAS results. This PRS derives from an eye colour GWA study that defines it quantitatively (i.e. 1 = blue or grey, 2 = green, 3 = hazel, and 4 = brown) completed by Lona-Durazo et al. using the CanPath cohort, which includes ∼5000 participants with European ancestry^23^. We estimated this PRS for each participant by extracting those genetic variants with a P-value<5·10^−8^ from the summary statistics and applying linear regression to the effects of these SNPs and the genotypes of our UKBB participants. We then included this PRS as covariate in an additional GWAS. Supplementary Fig. 7 demonstrates that the results of these GWAS are analogous to those of GWAS including both skin and hair colour.

QQ plots were generated using the R package qqman and ggplot2, and Manhattan plots and GWAS comparisons plots were generated using Matplotlib and seaborn libraries in python 3.

We completed a PheWAS to assess whether D_f_ loci have a significant effect in other traits. To this end, we searched D_f_ associated SNPs in GWASCatalog^25^ and GeneATLAS^24^. GWAScatalog contains hundreds of GWAS performed in different traits and populations and it constantly updates new GWAS to its database. GeneATLAS contains GWAS summary statistics for 778 UKBB traits and diseases using white European and white British participants from UKBB. These genetic variants have a P– value smaller than 5·10^−8^ on the trait in order to assume a strong association common to D_f_.

### GWAS and meta-analysis of Df loci across UKBB ancestries

We performed additional GWAS including UKBB participants with white European non-British (n_left_=4340 and n_right_=4288), Asian (n_left_=562 and n_right_=568) and African ancestries (n_left_=498 and n_right_=509) following the aforementioned model and procedure.

The meta-analysis was completed with those significant and independent SNPs from the D_f_ GWAS including white British participants. We extracted the summary statistics of these SNPs from the Asian, African, and white-European GWAS and performed a fixed-effect meta-analysis across UKBB ancestries. The meta-analysis and the forest plots were carried out with Meta package in R 4.0 software.

### Genetic correlation and heritability estimation

To investigate the shared genetic control between D_f_ and associated traits, we estimated their genome-wide genetic correlation. For this purpose, we obtained the GWAS summary statistics of traits of interest to our study from GeneATLAS and the eye colour study^23^. These calculations were computed with LD Score, a toolbox that estimates genetic correlation using GWAS summary statistics considering possible inflation caused by SNPs in linkage disequilibrium (LD). To ascertain the LD blocks within each variant, the software uses the 1000 Genomes panel as reference. Heatmaps were created with the genetic correlation estimate using the seaborn library in python 3.

The LD Score was also used to calculate the SNP heritability of both eyes’ D_f_. In this case, the software uses the reference map and the GWAS summary statistics to estimate the fraction of D_f_ variance explained by the SNPs’ additive effect.

### Mendelian Randomization

In order to infer the causality between the shared genetic basis of CAD, MI, hypertension and D_f_, we performed Mendelian Randomization analysis. For this procedure, we extracted the summary statistics of MI, hypertension, and CAD from GeneATLAS. We next selected for each cardiovascular condition separately those SNPs with a P-value<5·10^−08^, r^2^<0.001, MAF>0.01 which weren’t palindromic. The effect and the significance of these variants was also extracted from D_f_ GWAS summary statistics. We then estimated the causal effect of these genetic variants through different methods (inverse variance weighted regression, Egger’s regression, and Maximum likelihood) to analyse whether using different scenarios could better characterise the causality. This process was completed with TwoSamplesMR package in R 4.0. This package applies a quality control and a sensitivity analysis to evaluate the presence of palindromic SNPs, pleiotropy and heterogeneity which might influence the results of the study.

### Enrichment analysis

Functional and enrichment analysis were completed with FUMA to further analyse those metabolic pathways underlying significant D_f_ loci and its associated traits with a similar expression pattern. FUMA software not only characterise significant genetic variants from a GWAS study, but it also uses available GTEX data and available studies who implemented it to establish GO terms and traits with a similar expression profile^54,55^. We set the threshold of significant variants in a P-value<5·10^−06^ and a reference genomic panel of UKBB white Europeans.

### Development of MI predictive model

We used a subset of the UKBB data for MI model training and evaluation. We extracted white British UKBB participants who had good-quality images and a MI event after UKBB recruitment. MI events were defined in UKBB as a participant self-reporting MI at first repeated assessment visit [code 1075 from UKBB data field 20002] and MI hospitalizations identified using ICD10 codes [codes I21.1, I21.2, I21.3, I21.4, I21.9, I22, I22.0, I22.1, I22.8, I22.9,I23, I23.0,I23.1, I23.2, I23.3, I23.4, I23.5, I23.6, I23.8, I24.1 and I25.2 from UKBB data field 41204 and 41202]. The UKBB team previously validated this MI extraction algorithm and reported a minimum precision of 75%^56^. To define incident cases occurring after UKBB recruitment we used the date of the MI event [UKBB algorithmically defined MI event date from data field 42000] and the approximate time period when participants underwent to the ophthalmic examination, resulting in 526 incident cases. We randomly selected an equal number of age-matched participants (40-70 years) with good-quality images of both eyes no cardiovascular event within the CAD spectrum and no known risk factor (e.g., hypertension, and family history of heart disease).

Our MI predictive model uses age, sex, systolic blood pressure, smoking, BMI, a polygenic risk score for CAD and D_f_ of both eyes separately as features in a classification algorithm. We chose a random forest classifier algorithm allowing both non-linear associations between outcome and variables as well as inter-variable interaction in the model. Permutation-based feature importance scores^57^ were extracted in the modelling phase to assess the effect of each variable in the random forest construction using the *feature_importances_* function from the scikit-learn package. Given the influence of IQS on D_f_, we trained an additional model replacing D_f_ to D_f_ adjusted by IQS with to assess the existence of major differences in the model’s performance. We also tested whether introducing just one eye D_f_ in the model implied major differences in its performance.

To evaluate the performance of the predictive model, we reproduced SCORE with this MI dataset. SCORE uses age, sex, systolic blood pressure, and BMI as input variables for a logistic regression, with quantitative variables being discretized using healthcare guidelines^4^. We then assessed each model’s performance by using internal 10-fold cross validation and computing its AUC, precision, and recall. We used the same data partitions across SCORE and our MI models. A Wilcoxon signed-rank test was completed across all the trained models to evaluate the significance of the AUC differences.

We used Kaplan-Meier curves to assess the difference in survival rate difference between patients with high and low predicted MI probability, dichotomised at a probability of 0.5. This probability was obtained with our top-performing MI model. A Log-rank test was completed to evaluate the difference between these groups’ curves.

We investigated the sources of improvement of our MI model compared to the SCORE model through an ablation study. The model differs from SCORE in four key aspects:1) introducing D_f_, 2) the use of not-discretized quantitative variables, 3) using Random Forest instead of logistic regression, and 4) introducing PRS_CAD_. This ablation study consisted of assessing the performance of a modified version of SCORE through its AUC, recall and precision. These modifications included all the possible independent combinations across these alterations.

The code for this part of the study was written in Python 3.5.7 using the scitkit-learn, NumPy and Pandas packages and is available at https://github.com/Anavillaplana/MI_risk_prediction. ROC curves were plotted using the predicted MI probability from each model using the ROCurve plot package in R 4.0. Both Kaplan-Meier curves and the Log-rank test were completed with the lifelines Python package.

### Estimating CAD polygenic risk score

PRS_CAD_ derives from the CARDIoGRAMplusC4D Consortium^15^ which is one of the largest completed CAD meta-analysis. This study doesn’t include UKBB data, but it’s developed with multiple CAD databases with different ancestries to better characterise the genetic control of this outcome. We estimated PRS_CAD_ for each participant in the MI dataset by using PRSice-2 software^58^, the summary statistics of the meta-analysis, and the genotypes of this MI dataset. We then included this PRS as variable in our MI predictive model.

## Data Availability

All data produced in the present study are available upon reasonable request to the authors

## Acknowledgements

This research has been conducted using the UK Biobank Resource under project 788. This work is supported by the Medical Research Council grant (MR/N013166/1) to A.V.V; the Roslin Institute Strategic Programme Grant from the BBSRC (BBS/E/D/30002275 and BBS/E/D/30002276) to A.T; Health Data Research UK grants (references HDR-9004 and HDR-9003) to A.T; the Engineering and Physical Sciences Research Council (EPSRC) grant (EP/R029598/1, EP/T008806/1) to M.O.B; Fondation Leducq grant (17 CVD 03) to M.O.B, the European Union’s Horizon 2020 research and innovation programme under grant agreement No 801423 to M.O.B; Diabetes UK grant (20/0006221) to M.O.B; Fight for Sight grant (5137/5138) to M.O.B; and British Heart Foundation and The Alan Turing Institute (which receives core funding under the EPSRC grant EP/N510129/1) as part of the Cardiovascular Data Science Awards Round 2 (SP/19/9/34812) to M.O.B. Support from NHS Lothian R&D and the Edinburgh Clinical Research Facility is acknowledged.

