## Supplementary Material for "Decreased retinal vascular complexity is an early biomarker of MI supported by a shared genetic control"

SUPPLEMENTARY MATERIALS

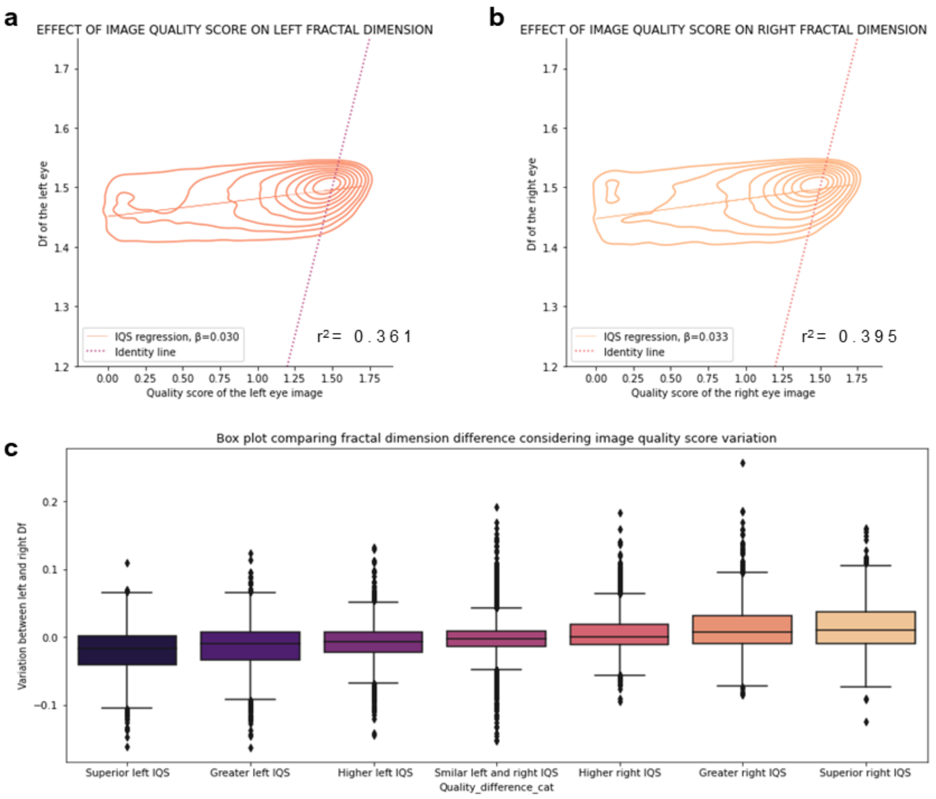


**Supplementary Figure 1: Effect between image quality score (IQS) and fractal dimension.** The contour plot evidences the joint distribution of **a** left and **b** right fractal dimension and image quality score. **C** The box plot illustrates the interocular fractal dimension difference at multiple IQS variation cases.

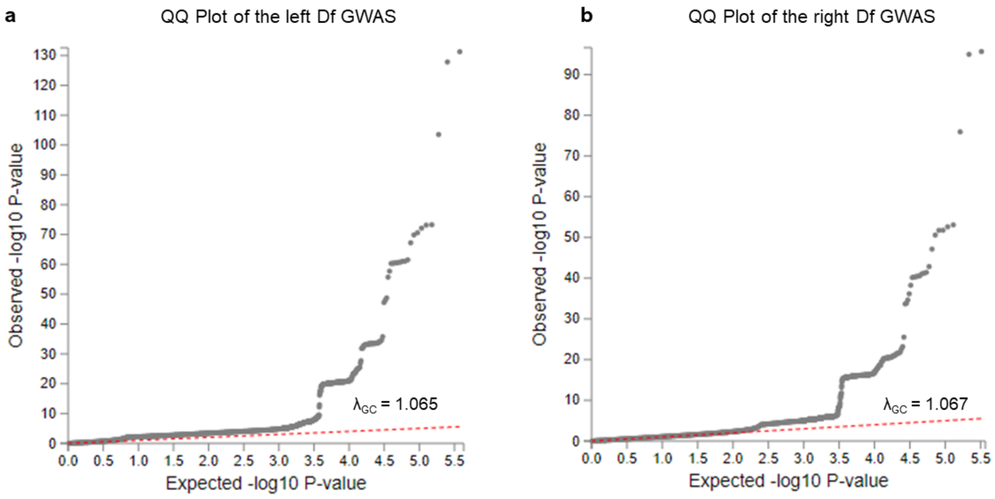


**Supplementary Figure 2: QQ plots for the GWAS.** These plots illustrate the expected vs observed -Log(P-value) comparison obtained in **a** the left and **b** right Df GWAS.

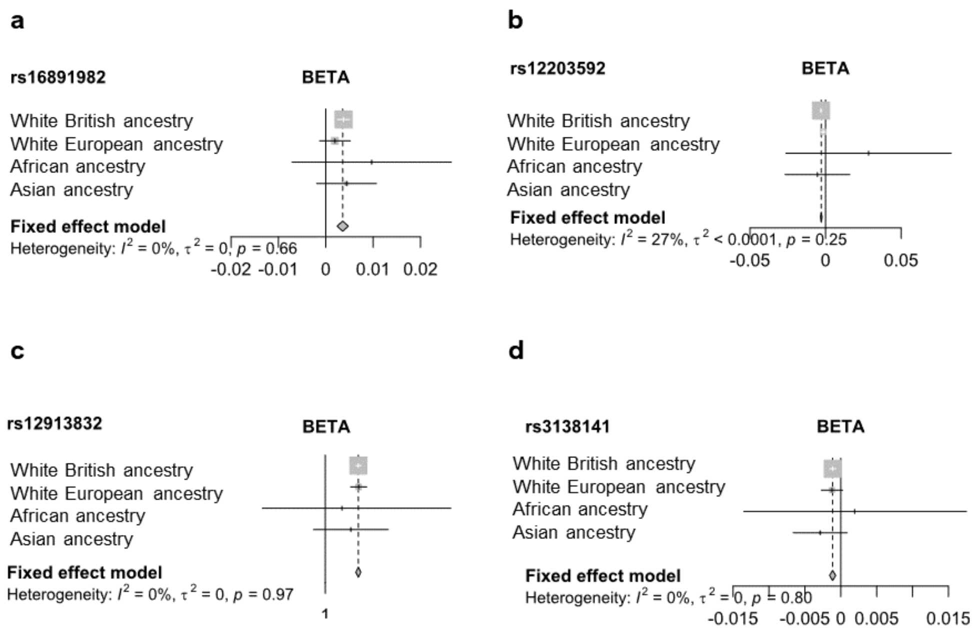


**Supplementary Figure 3: Forest plots of Df-associated SNPs.** These illustrate the effect of significant genetic variants (**a** rs16891982 **b** rs12203592 **c** rs12913832 **d** rs3138141) across UKB ancestries.

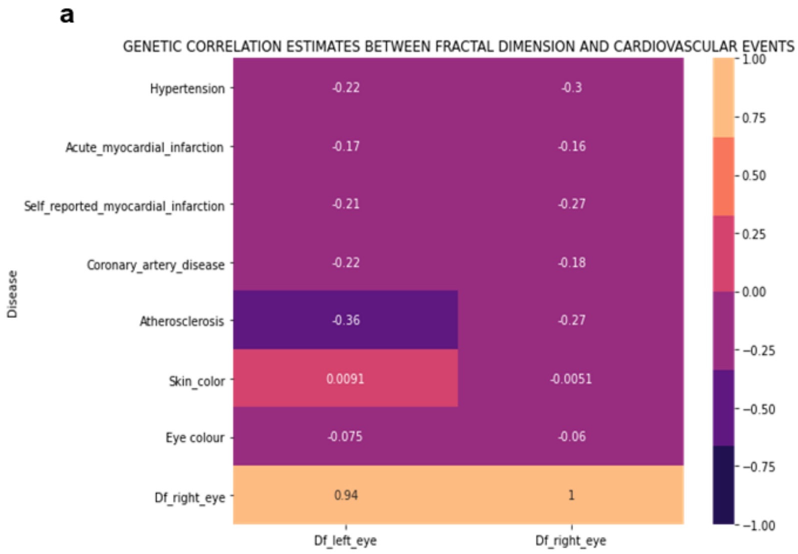


**b**

| Correlated trait | LEFT EYE | | | RIGHT EYE | | |
|---|---|---|---|---|---|---|
| | $r_g^2$ | SE | P value | $r_g^2$ | SE | P value |
| Hypertension | -0.2229 | 0.0534 | 3.0261e-05 | -0.3020 | 0. 0659 | 4.52e-06 |
| Acute myocardial infarction | -0.1717 | 0.0809 | 0.0801 | -0.1585 | 0.1075 | 0.0308 |
| Self reported acute myocardial infarction | -0.2071 | 0. 0754 | 0.006 | -0.2663 | 0. 0982 | 0.0067 |
| Coronary artery disease | -0.2214 | 0.0591 | 1.7854e-04 | -0.1776 | 0. 0795 | 0.025 |
| Atherosclerosis | -0.3585 | 0.1819 | 0.084 | -0.2668 | 0.2100 | 0.051 |
| Skin colour | 0.0091 | 0.0536 | 0.8647 | -0.0051 | 0. 0568 | 0.9282 |
| Eye colour | -0.0751 | 0.0334 | 0.6372 | -0.0598 | 0.0271 | 0.7658 |
| Right Fractal dimension | 0.9468 | 0.0962 | 1.75E-26 | - | | - |

**Supplementary Figure 4: Genetic correlations between fractal dimension and associated traits. a** Heatmap illustrating the direction and percentage of shared genomic regions, also indicated by the number. **b** Table describing genetic correlation estimates and its P-value

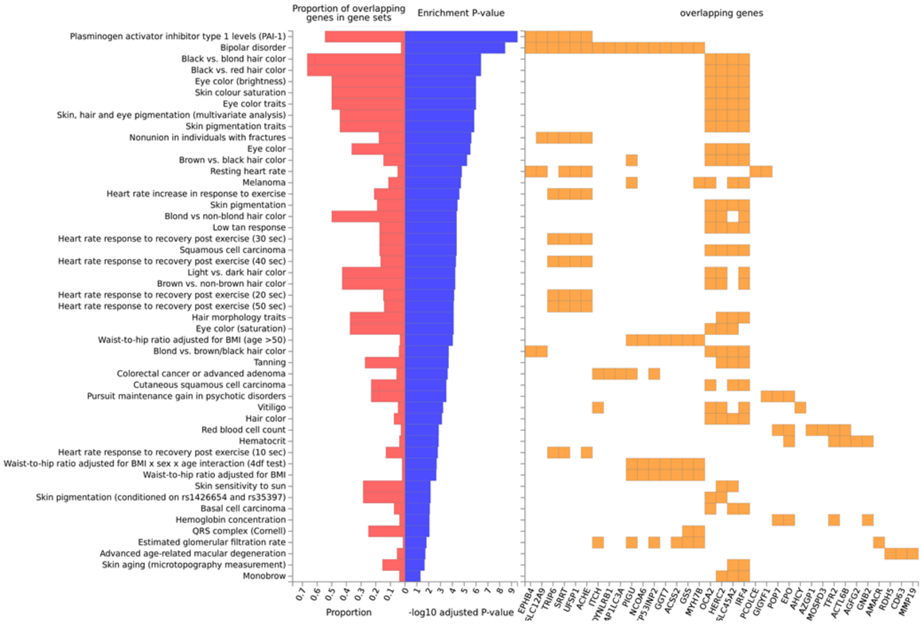


**Supplementary Figure 5: Traits and pathways sharing significant genetic effects with fractal.**

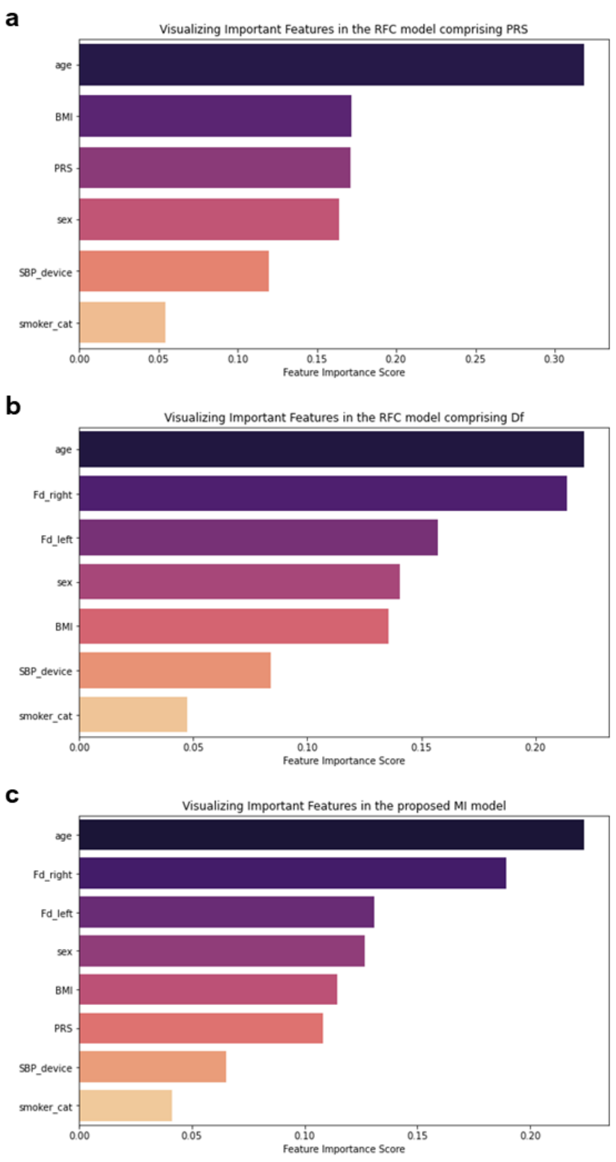


**Supplementary Figure 6: Feature importance score of the MI models based on a random forest classifier.** SBP device: Systolic blood pressure measured at baseline UKB assessment using their automatic device. PRS: CAD polygenic risk score based on CARdioGRAM consortium. BMI: basal muscular index. Fd_left and Fd_right: measures of left and right fractal dimension, respectively. Smoker cat: participants who are current smokers at baseline examination.

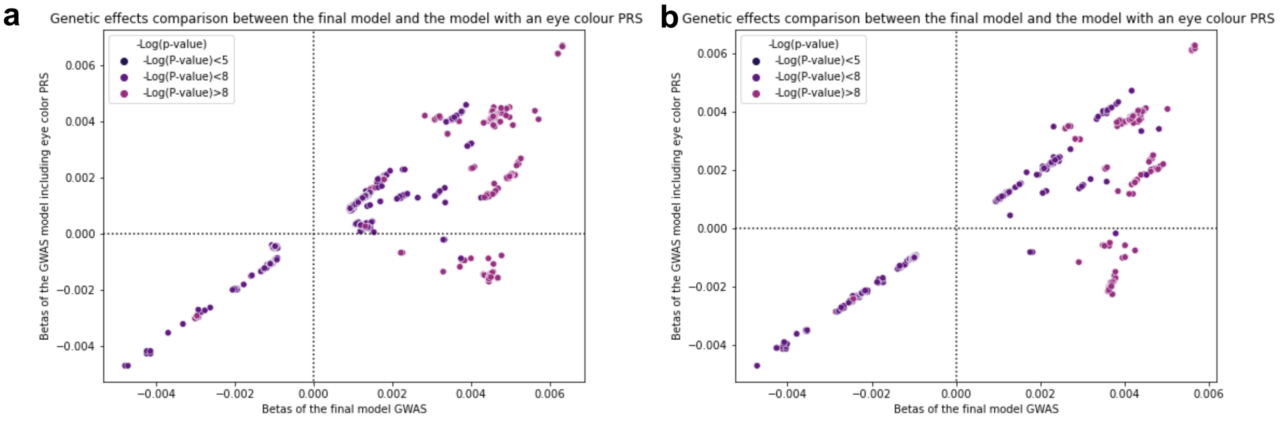

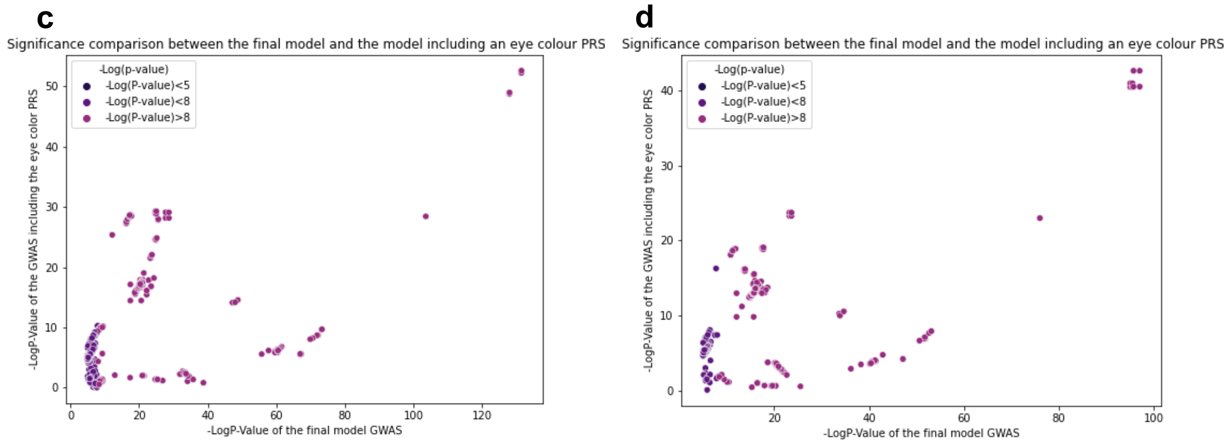


**Supplementary Figure 7: Left and right eye GWAS results comparisons between the final model and this model including an eye colour PRS. a** Left and **b** right eye scatterplot illustrating the SNP's effect comparison between the final GWAS model and the GWAS including the pigmentation and the eye colour PRS. **c** Left and **d** right eye scatterplot representing the -Log(P-value) comparison among these two models. SNPs with a –Log10(P)=4.5 are only included in these figures for clarity.

**Supplementary Table 1: Summary statistics of UKB traits.** The table includes the linear regression effect, its standard deviation, P-value, Pearson correlation and its P-value. This table is within the excel spreadsheet.

**Supplementary Table 2: GWAS summary statistics.** Includes the MAF, SNP effect, SD and -Log(P-value) of all significant SNPs for both eyes. The nearest gene, its association with ocular or non-ocular traits it's also included.

| SNP | CHR | MAF | LEFT EYE | | | RIGHT EYE | | | Nearest gene | Ocular association | Non-ocular association |
|---|---|---|---|---|---|---|---|---|---|---|---|
| | | | BETA | SD | -Log (P-value) | BETA | SD | -Log (P-value) | | | |
| rs1709657 | 3 | 0.23 | 9.42E-04 | 2.40E-04 | 4.15 | 1.13E-03 | 2.49E-04 | 5.33 | *EIF2B5* | | Inflammation and platelet count |
| rs16891982 | 5 | 0.024 | 3.75E-03 | 6.59E-04 | 7.93 | 3.53E-03 | 6.94E-04 | 6.46 | *SLC45A2* | | Skin, hair and eye colour and pigmentation disease |
| rs12203592 | 6 | 0.22 | -2.31E-03 | 2.68E-04 | 28.67 | -2.85E-03 | 2.80E-04 | 23.62 | *IRF4* | Refractive error | Skin, hair colour, eye colour and disease and lymphocyte and leukocyte count. |
| rs80308281 | 7 | 0.005 | -6.79E-03 | 1.46E-03 | 6.73 | -7.19E-03 | 1.36E-03 | 6.88 | *SLC12A9* | Choroid and retinal disease and retinal detachment | Skin colour, hair colour and disease, mean arterial pressure and resting heart rate |
| rs56108400 | 12 | 0.24 | -1.07E-03 | 2.37E-04 | 5.17 | -1.06E-03 | 2.48E-04 | 5.72 | *RDH5/ ORMDL2* | Macular thickness, AMD, retinitis pigmentosa, disorders of the lens, cataract, ocular muscle, myopia and retinal detachment | |
| rs12913832 | 15 | 0.22 | 6.34E-03 | 2.58E-04 | 131.28 | 5.65E-03 | 2.71E-04 | 96.97 | *HERC2* | Cataract, retinal arterial and venular width and tortuosity, visual acuity, AMD and IOP | Pulse pressure and hair, skin and eye colour |
| rs916977 | 15 | 0.15 | 5.12E-03 | 2.94E-04 | 66.93 | 4.48E-03 | 3.08E-04 | 47.13 | *OCA2* | Cataract, IOP, lens disorders and Glaucoma | Hair, skin, and eye colour |
| rs4911429 | 20 | 0.31 | -5.49E-04 | 2.23E-04 | 5.54 | -1.33E.03 | 2.33E-04 | 7.98 | *MAPILC3A* | | Skin colour, platelet and reticulocyte count |
| rs73226964 | 21 | 0.02 | -3.21E-03 | 7.48E-04 | 4.63 | -4.00E-03 | 7.75E-04 | 6.63 | *AGPAT3* | | RBC count, eosinophil percentage, GFR diabetes miellitus, and disorders of the arteries. |

**Supplementary Table 3: Comparison between Zekavat et al. GWAS and this study.** Includes reported genetic variants

| | | | Zekava et al[56] | | This study | |
|---|---|---|---|---|---|---|
| **SNP** | **MAF** | **NEAREST GENE** | **BETA** | **-Log (P-value)** | **BETA** | **-Log (P-value)** |
| rs12203592 | 0.22 | *IRF4* | -0.05 | 13 | -2.85E-03 | 23.62 |
| rs12913832 | 0.22 | *HERC2* | 0.15 | 78 | 5.65E-03 | 96.97 |
| rs7164220 | 0.15 | *OCA2* | 0.11 | 65 | 4.61E-03 | 51.76 |

**Supplementary Table 4: Mendelian randomization results.** Includes the heterogeneity and pleiotropy test as well as the statistic and P-value of the MR methods used for both cardiovascular outcomes in both eyes.

| | Left Df | | | | | | Right Df | | | | | |
|---|---|---|---|---|---|---|---|---|---|---|---|---|
| Cardiovascular event | Heterogeneity | Horizontal Pleiotropy | Inverse variance weighted | P-value | Maximum likelihood | P-value | Heterogeneity | Horizontal Pleiotropy | Inverse variance weighted | P-value | Maximum likelihood | P-value |
| Myocardial infarction | 0.367 | 0.006 | 0.080 ±0.101 | 0.429 | 0.080 ±0.102 | 0.427 | 0.394 | 0.007 | 0.095 ±0.104 | 0.358 | 0.096 ±0.104 | 0.357 |
| Mild Ischaemic heart disease | 0.552 | 0.033 | -0.064 ±0.122 | 0.600 | -0.064 ±0.125 | 0.610 | 0.619 | 0.029 | -0.074 ±0.049 | 0.533 | -0.074 ±0.052 | 0.551 |

**Supplementary Table 5: Demographics of the MI cases and controls.** This table describes for cases and controls the centrality and dispersion of the epidemiological variables included in the MI predictive model. P-value* refers to the T-test completed among these two groups to estimate its difference.

| Variable | MI cases | Control cases | P-Value* |
|---|---|---|---|
| Age (years) | 57.31±6.47 | 54.21±7.84 | 1.076e-39 |
| Sex (N.Females/N.Males) | 122/403 | 298/227 | - |
| BMI | 28.54±4.63 | 26.42±4.39 | 5.52e-14 |
| SBP (mmHg) | 142.03±20.20 | 135.37±18.01 | 2.21e-8 |
| Current smokers | 69 | 28 | - |
| Right $D_f$ | 1.485±0.03 | 1.5±0.075 | 3.33e-15 |
| Left $D_f$ | 1.485±0.03 | 1.494±0.036 | 1.97e-08 |
| $PRS_{CAD}$ | 3.58±0.32 | 3.41±0.24 | 1.94e-06 |

**Supplementary Table 6: Wilcoxon signed-rank test across the examined MI models.** This table includes the Wilcoxon signed-rank sum test and the P-Value of all the comparisons between the models we trained in this study.

| Models comparison | Wilcoxon signed rank sum test | P-Value* |
|---|---|---|
| SCORE VS RFC with $D_f$ and $PRS_{CAD}$ | 1.0 | 0.0039 |
| SCORE VS RFC with $D_f$ | 1.0 | 0.0039 |
| SCORE VS RFC with $PRS_{CAD}$ | 6.0 | 0.027 |
| RFC with $D_f$ and $PRS_{CAD}$ VS RFC with $PRS_{CAD}$ | 7.0 | 0.037 |
| RFC with $D_f$ and $PRS_{CAD}$ VS RFC with $D_f$ | 25.0 | 0.846 |
| RFC with $PRS_{CAD}$ VS RFC with $D_f$ | 7.0 | 0.037 |

In our ablation study we found three central elements ascertaining the superior accuracy of our model on distinguishing personalised MI risk in UKB. Firstly, the usage of continuous variables. Those models following this premise yield a greater performance than SCORE and its derivates (i.e., SCORE + $D_f$). We further investigated whether this situation was reproduced in RFC-based classifiers. Our results show that these models achieve a higher AUC when compared with the ones that introduce age, BMI and SBP as discrete variables. Secondly, we observe that the presence of $D_f$ and $PRS_{CAD}$ in the predictive model significantly improves its performance, regardless of the classifier's algorithm. Finally, RFC-based models yield higher AUC, precision, and recall when compared with SCORE and all the completed transformations, implying that a non-linear algorithm benefit individual MI prediction. Amongst these RFC classifiers, the one including both aforementioned elements achieves the greatest performance, followed by a similar model excluding $PRS_{CAD}$.

**Supplementary Table 7: Ablation study of the MI models.** These tables include the precision, recall and AUC for the variations in the model.* AUC estimates significantly different (Wilcoxson signed-rank test P-value<0.005) from the one obtained with the SCORE model.** AUC estimates significantly different (Wilcoxson signed-rank test P-value<0.005) from the one obtained with the SCORE model and the one from our final model.

| | **MI** | | |
|---|---|---|---|
| Model | Precision | Recall | AUC |
| SCORE model[16] | 0.705 ±0.00096 | 0.729 ±0.0019 | 0.711 ±0.0008 |
| SCORE model and Df | 0.705 ±0.00083 | 0.731 ±0.0021 | 0.711 ±0.00086 |
| SCORE model and $PRS_{CAD}$ | 0.721 ±0.001 | 0.739 ±0.0016 | 0.723 ±0.0011 |
| SCORE model and Df + $PRS_{CAD}$ | 0.718 ±0.0012 | 0.737 ±0.0019 | 0.727 ±0.0009 |
| SCORE model using continuous variables | 0.732 ±0.0014 | 0.752 ±0.0024 | 0.737 ±0.0013 |
| SCORE model with continuous variables and Df** | 0.728 ±0.0014 | 0.754 ±0.0023 | 0.735 ±0.0013 |
| SCORE model with continuous variables and $PRS_{CAD}$** | 0.753 ±0.00096 | 0.768±0.0013 | 0.747 ±0.00083 |
| SCORE model with continuous variables and Df + $PRS_{CAD}$* | 0.752 ±0.001 | 0.775±0.0013 | 0.759 ±0.00086 |
| SCORE model using a RFC | 0.711 ±0.0013 | 0.745 ±0.0007 | 0.719 ±0.001 |
| SCORE model using a RFC and Df* | 0.758 ±0.0013 | 0.765 ±0.0016 | 0.748 ±0.0013 |
| SCORE model using a RFC and $PRS_{CAD}$ | 0.727 ±0.0012 | 0.737 ±0.0022 | 0.728 ±0.0011 |
| SCORE model using a RFC and Df + $PRS_{CAD}$** | 0.745 ±0.001 | 0.781 ±0.0014 | 0.752 ±0.0009 |
| SCORE model using continuous variables and RFC** | 0.733 ±0.0013 | 0.750 ±0.0024 | 0.738 ±0.0013 |
| SCORE model using continuous variables, Df and RFC* | 0.756 ±0.0008 | 0.778±0.0013 | 0.763 ±0.0011 |
| SCORE model using continuous variables, $PRS_{CAD}$ and RFC** | 0.735 ±0.0010 | 0.756 ±0.0021 | 0.741±0.0012 |
| SCORE model using continuous variables, Df, $PRS_{CAD}$ and RFC* | **0.763±0.0016** | **0.788 ±0.0014** | **0.770 ±0.0013** |

**Supplementary Table 8: Additional MI models performance.** These tables include the precision, recall and AUC for the variations in th0se supplementary MI models.

| | MI | | |
|---|---|---|---|
| Model | Precision | Recall | AUC |
| Random Forest including left Df | 0.756±0.001 | 0.773±0.002 | 0.761±0.001 |
| Random Forest including left Df and $PRS_{CAD}$ | 0.759±0.001 | 0.793±0.001 | 0.774±0.001 |
| Random Forest including right Df | 0.759±0.001 | 0.778±0.002 | 0.765±0.001 |
| Random Forest including right Df and $PRS_{CAD}$ | 0.756±0.001 | 0.787±0.001 | 0.772±0.001 |
| Random Forest including quality-adjusted Df | 0.766±0.001 | 0.784±0.002 | 0.771±0.001 |
| Random Forest including quality-adjusted Df and $PRS_{CAD}$ | 0.768±0.001 | 0.796±0.001 | 0.774±0.001 |
